# 3D-DXA Cortical and Trabecular Parameters; Agreement and Precision Between GE Healthcare Prodigy and iDXA Densitometers

**DOI:** 10.64898/2026.03.04.26347524

**Authors:** Diane Krueger, Neil Binkley, Miguel Madeira, Zhaojing Chen, Silvana Di Gregorio, Luis Del Río, Ludovic Humbert

**Author notes:** **Corresponding author:** Diane Krueger.

## Abstract

3D-DXA reconstructs DXA hip scans to 3-dimensional images allowing measurement of trabecular and cortical bone parameters. Given the higher image quality of GE Healthcare iDXA than GE Healthcare Prodigy, it could be hypothesized that the reconstruction might differ, thereby affecting 3D-DXA results. The aim of the study was to assess agreement and precision of 3D-DXA cortical and trabecular femur parameters between Prodigy and iDXA densitometers in adult subjects. The study cohort was composed of 391 men and women recruited from 3 clinical centers (USA and Brazil). All subjects were scanned on either Prodigy or iDXA scanners. Short-term precision was assessed on two Prodigy and two iDXA densitometers. 3D-DXA analyses were performed using 3D-Shaper software version 2.14. Agreement between densitometers was assessed by regression and Bland-Altman analyses. Short-term precision was determined following International Society for Clinical Densitometry recommendations. Strong agreements for 3D-DXA parameters were obtained between devices regardless of the center or the DXA device model (all R^2^ > 0.96). Bland-Altman analyses demonstrated statistically (p < 0.05), but not clinically, significant difference between both aBMD and 3D-DXA measurements obtained using Prodigy and iDXA scanners. Short-term precision of areal BMD and 3D-DXA parameters was similar between densitometers. This study demonstrated excellent 3D-DXA measurement agreement and similar precision between iDXA and Prodigy densitometers. These data provide evidence that no adjustments are required when using 3D-Shaper software on iDXA or Prodigy instruments.

**Mini Abstract:** We assessed agreement and precision of 3D-DXA parameters between GE Healthcare Prodigy and iDXA densitometers in adults. Strong agreement was observed between devices, and short-term precision was comparable. Findings indicate that no adjustment is needed when using 3D-DXA with GE Healthcare densitometers.

## Introduction

Dual-energy X-ray absorptiometry (DXA) is the current gold standard imaging technique for the clinical diagnosis of osteoporosis based on bone mineral density (BMD) measurement. Over the last 20 years, new technologies have been developed to enhance the broad applicability of DXA [1–6]. Among them, 3D modelling methods (3D-DXA) allow clinicians access to proximal femur information equivalent to that provided by Quantitative Computed Tomography (QCT) [5, 6]. Those approaches register a 3D statistical model onto a standard hip DXA scan to provide a subject-specific model of the proximal femur allowing assessment of trabecular and cortical bone compartments.

While short term precision errors and measurement agreement between DXA systems have been studied extensively [7–10], sparse data exists for 3D-DXA techniques [11]. Specifically, precision error and least significant change (LSC) of 3D-DXA measurements have only been evaluated for Lunar iDXA (GE Healthcare, Madison, WI, USA) and Discovery W (Hologic, Marlborough, MA, USA) DXA scanners [11]. In that same study, authors also observed the trend assessment intervals (TAI) in postmenopausal women for both areal BMD and 3D-DXA measurements at the proximal femur were similar [11]. However, no data is available for 3D-DXA measurement agreement between DXA systems.

The aim of the present study is to evaluate both agreement and precision of 3D-DXA cortical and trabecular parameters between GE Healthcare Prodigy and iDXA densitometers in adults.

## Material and methods

### Subjects

The study cohort was composed of 391 men and women recruited from 3 clinical centers. For all centers, subjects referred for bone health assessment were recruited considering the following inclusion and exclusion criteria: age ≥ 20 years, no surgical bone implant or anatomical abnormalities that can impact DXA measurements. Pregnant women and those with a fracture within the last 12 months were excluded. More details about subject eligibility for each center can be found elsewhere [8, 12]. Among them, 330 (145 men and 185 women) were included from the University of Wisconsin (***UW***, Madison, WI, US). Thiry young adults (10 men and 20 women) were recruited from California State University, San Bernardino (***CSUSB***, San Bernardino, CA, US). Thirty-one (1 men and 30 women) were recruited from Federal University of Rio de Janeiro (***FURJ***, Rio de Janeiro, Brazil). The mean age of the included subjects were 52 years (±18.5 years), 23 years (±1.8 years) and 53 years (±10.8 years) for the UW, CSUSB and FURJ respectively.

All subjects provided signed informed consent to participate in the respective studies conducted in accordance with the principles in the Declaration of Helsinki with approval from their university’s research ethics committee.

### DXA Measurements

All subjects were scanned following manufacturer recommendations to measure areal bone mineral density (aBMD) with the following DXA devices: a Prodigy and iDXA scanner at the UW and CSUSB, two Prodigy devices were used at FURJ. Each subject was scanned on the two scanners at the respective sites using enCORE software as specified in Table 1. Left hip was assessed at UW, right hip was assessed at FURJ while both hips were assessed at CSUSB. Finally, short-term precision studies were performed on both the Prodigy and iDXA scanners at UW in a sub-cohort of 31 women, on both Prodigy devices at FURJ and on the iDXA scanner at CSUSB. Duplicate hip scans with complete repositioning between acquisitions were acquired (two acquisitions per DXA device).

**Table 1.**
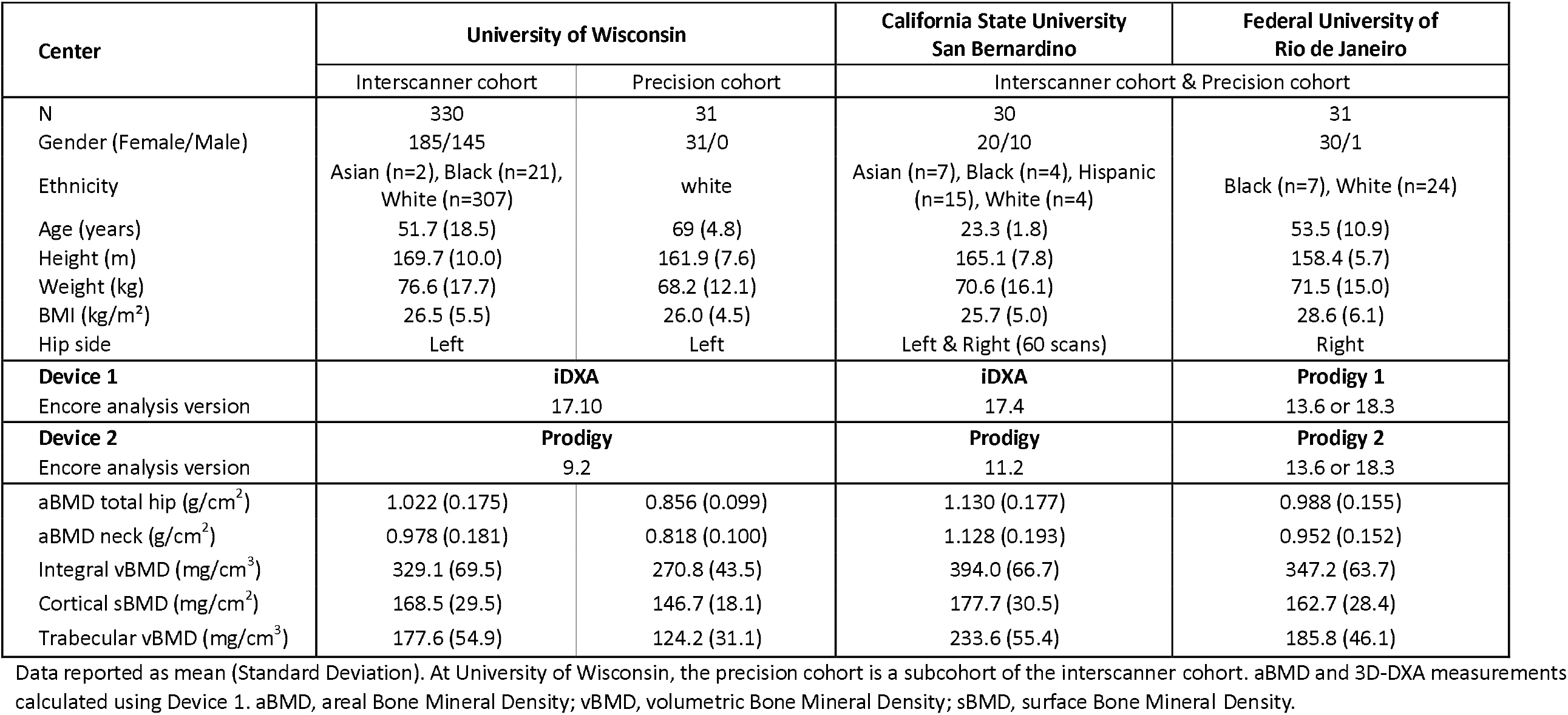
Subject demographics, DXA device and software, aBMD and 3D-DXA measurements by center.

| Center | University of Wisconsin |  | California State University<br>San Bernardino | Federal University of<br>Rio de Janeiro |
| --- | --- | --- | --- | --- |
|  | Interscanner cohort | Precision cohort | Interscanner cohort & Precision cohort |  |
| N | 330 | 31 | 30 | 31 |
| Gender (Female/Male) | 185/145 | 31/0 | 20/10 | 30/1 |
| Ethnicity | Asian (n=2), Black (n=21),<br>White (n=307) | white | Asian (n=7), Black (n=4), Hispanic<br>(n=15), White (n=4) | Black (n=7), White (n=24) |
| Age (years) | 51.7 (18.5) | 69 (4.8) | 23.3 (1.8) | 53.5 (10.9) |
| Height (m) | 169.7 (10.0) | 161.9 (7.6) | 165.1 (7.8) | 158.4 (5.7) |
| Weight (kg) | 76.6 (17.7) | 68.2 (12.1) | 70.6 (16.1) | 71.5 (15.0) |
| BMI (kg/m<sup>2</sup>) | 26.5 (5.5) | 26.0 (4.5) | 25.7 (5.0) | 28.6 (6.1) |
| Hip side | Left | Left | Left & Right (60 scans) | Right |
| **Device 1** | **iDXA** |  | **iDXA** | **Prodigy 1** |
| Encore analysis version | 17.10 |  | 17.4 | 13.6 or 18.3 |
| **Device 2** | **Prodigy** |  | **Prodigy** | **Prodigy 2** |
| Encore analysis version | 9.2 |  | 11.2 | 13.6 or 18.3 |
| aBMD total hip (g/cm<sup>2</sup>) | 1.022 (0.175) | 0.856 (0.099) | 1.130 (0.177) | 0.988 (0.155) |
| aBMD neck (g/cm<sup>2</sup>) | 0.978 (0.181) | 0.818 (0.100) | 1.128 (0.193) | 0.952 (0.152) |
| Integral vBMD (mg/cm<sup>3</sup>) | 329.1 (69.5) | 270.8 (43.5) | 394.0 (66.7) | 347.2 (63.7) |
| Cortical sBMD (mg/cm<sup>2</sup>) | 168.5 (29.5) | 146.7 (18.1) | 177.7 (30.5) | 162.7 (28.4) |
| Trabecular vBMD (mg/cm<sup>3</sup>) | 177.6 (54.9) | 124.2 (31.1) | 233.6 (55.4) | 185.8 (46.1) |

Data reported as mean (Standard Deviation). At University of Wisconsin, the precision cohort is a subcohort of the interscanner cohort. aBMD and 3D-DXA measurements calculated using Device 1. aBMD, areal Bone Mineral Density; vBMD, volumetric Bone Mineral Density; sBMD, surface Bone Mineral Density.

Device calibrations were checked and passed on a daily basis before each scanning session, using the GE Healthcare calibration block.

### 3D-DXA analysis

3D-DXA analyses were performed using 3D-Shaper® software (v2.14, 3D-Shaper Medical, Barcelona, Spain). Details of the 3D-Shaper methodology can be found elsewhere [5]. In short, 3D-Shaper software assesses the proximal femur in 3D from a standard hip DXA scan and provides clinicians with advanced characterization of the cortical and trabecular structures. The 3D-Shaper algorithm registers a statistical shape and density 3D model onto the hip DXA scans of the patient to create a patient specific 3D femur. The cortex is subsequently analyzed by fitting a mathematical function to the density profile computed along the normal vector at each node of the proximal femur surface mesh [13]. Three measurements are performed to characterize the 3D femur:

- Integral volumetric BMD (Integral vBMD, expressed in mg/cm^3^) is calculated as the mean density of the integral (i.e., cortical and trabecular bones at total hip region) compartment.
- Cortical surface BMD (sBMD; expressed in mg/cm^2^) is calculated, at total hip region, as the product of cortical vBMD and cortical thickness.
- Trabecular volumetric BMD (trabecular vBMD, expressed in mg/cm^3^) is calculated as the average density of the trabecular compartment at total hip region.

### Statistical Analysis

Statistical analyses were carried out using Microsoft Excel 2019 (Microsoft Corporation, Redmond, WA, USA). Demographics and baseline characteristics were summarized and presented by center. Baseline subject characteristics were reported for quantitative variables as mean ± SD or median (first quartile-third quartile) and for categorial variables as number or percentage. Agreement between densitometers was assessed using Deming regression (assuming equal error variance) and Bland-Altman analysis. From the Bland-Altman analysis, the following limits of agreement parameters were extracted: interval of agreement at 95% and the width of the interval. The width of the interval provides an indication of the magnitude of the differences that are to be expected between pairs of measurements. Difference between measurements across DXA models (paired samples from a center) were evaluated using a F-Test. Precision error was obtained in compliance with the ISCD (International Society for Clinical Densitometry) recommendations [14]. From the duplicate scans, in vivo short-term precision errors for aBMD and 3D measurements were evaluated using the root-mean-square standard deviation and root-mean-square coefficient of variation. The least significant change (LSC) for the 95% confidence level was defined as 2.77 times the precision error. An overall two-sided significance level of 0.05 was used.

## Results

Subject characteristics, DXA device, Encore® software (GE Healthcare) versions used to analyze the DXA scans, aBMD and 3D-DXA measurements are presented in Table 1.

### Inter-scanner agreements

#### Areal Bone Mineral Density

The comparison between Prodigy and iDXA scanners showed excellent agreements of aBMD (total hip and femoral neck) regardless of the center (Table 2). Measurements were highly correlated as characterized by a R^2^ ranging from 0.97 to 0.99 (Table 2, Figure 1,2 and 3). Per center, the aBMD bias were similar at total hip and femoral neck (Table 2, Figure 1,2 and 3). Although similar for both regions of interest, numerically smaller biases were observed at the UW (−0.001 and −0.006 g/cm^2^; for aBMD at total hip and femur neck respectively) and at FURJ (−0.005 and −0.002 g/cm^2^ for aBMD at total hip and femur neck respectively), compared to CSUSB (−0.016 and −0.021 g/cm^2^ for aBMD at total hip and femur neck respectively). These biases were significant for aBMD at femoral neck between Prodigy and iDXA at the UW (p<0.001) and at CSUSB (p<0.001) while no significant bias was observed at the total hip at the UW (p=0.41). In addition, significant biases were obtained at FURJ at the total hip but not femoral neck (p=0.048 and p=0.73 respectively). Similar RMSE were obtained across the centers (all F-test not significant, data not shown) for aBMD at total hip or at femoral neck. Limits of agreement are presented in Table 2. The intervals were similar for measurements from Prodigy and iDXA devices regardless of the center, indicating good agreement for measurements between the two scanners. A narrower interval was obtained for total hip aBMD measurements of Prodigy versus Prodigy (0.056 g/cm^2^ at FURJ), compared to Prodigy versus iDXA (0.074 g/cm^2^ at UW and 0.106 g/cm^2^ at CSUSB). Similar Intervals were observed for aBMD at femoral neck at UW and CSUSB (0.080 and 0.074 g/cm^2^ respectively) while a slightly higher interval was obtained at FURJ (0.102 g/cm^2^).

**Table 2.** Agreement between devices by center for aBMD and 3D-DXA measurements.

| Center | Devices | N | Regression or Bland-Altman analysis | aBMD total hip (g/cm<sup>2</sup>) | aBMD neck (g/cm<sup>2</sup>) | Integral vBMD (mg/cm<sup>3</sup>) | Cortical sBMD (mg/cm<sup>2</sup>) | Trabecular vBMD (mg/cm<sup>3</sup>) |
| --- | --- | --- | --- | --- | --- | --- | --- | --- |
| University of Wisconsin | Prodigy versus iDXA | 330 | R<sup>2</sup> | 0.99 | 0.99 | 0.99 | 0.98 | 0.98 |
|  |  |  | Bias | -0.001 | -0.006 | -2.4 | -0.9 | -2.7 |
|  |  |  | RMSE | 0.015 | 0.023 | 8.7 | 4.5 | 8.2 |
|  |  |  | Slope | 1.000 | 0.998 | 1.002 | 0.976 | 0.972 |
|  |  |  | Intercept | -0.001 | 0.006 | -2.937 | 3.198 | 2.28 |
|  |  |  | Interval of agreement 95% | [-0.038 : 0.036] | [-0.046 : 0.034] | [-15.1 : 10.3] | [-8.5 : 6.7] | [-13.8 : 8.4] |
|  |  |  | Limit of Agreement Interval width | 0.074 | 0.080 | 25.4 | 15.2 | 22.2 |
|  |  |  | p | 0.41 | <0.001 | <0.001 | <0.001 | <0.001 |
| Federal University of Rio de Janeiro | Prodigy 1 versus Prodigy 2 | 31 | R<sup>2</sup> | 0.99 | 0.97 | 0.99 | 0.98 | 0.99 |
|  |  |  | Bias | -0.005 | -0.002 | 1.2 | 0.1 | 0.4 |
|  |  |  | RMSE | 0.015 | 0.027 | 6.5 | 3.7 | 5.5 |
|  |  |  | Slope | 0.967 | 1.022 | 0.991 | 1.001 | 0.980 |
|  |  |  | Intercept | 0.027 | -0.023 | 4.182 | -0.060 | 4.093 |
|  |  |  | Interval of agreement 95% | [-0.033 : 0.023] | [-0.053 : 0.049] | [-13.8 : 16.2] | [-7.1 : 7.3] | [-10.3 : 11.1] |
|  |  |  | Limit of Agreement Interval width | 0.056 | 0.102 | 30.0 | 14.4 | 21.4 |
|  |  |  | p | 0.048 | 0.73 | 0.32 | 0.83 | 0.72 |
| California State University San Bernardino | Prodigy versus iDXA | 30 | R<sup>2</sup> | 0.99 | 0.98 | 0.99 | 0.99 | 0.98 |
|  |  |  | Bias | 0.016 | 0.021 | 3.0 | 0.8 | -0.8 |
|  |  |  | RMSE | 0.023 | 0.037 | 8.7 | 3.7 | 6.9 |
|  |  |  | Slope | 1.053 | 1.054 | 1.038 | 1.007 | 1.016 |
|  |  |  | Intercept | -0.043 | -0.038 | -11.644 | -0.342 | -4.524 |
|  |  |  | Interval of agreement 95% | [-0.037 : 0.069] | [-0.016 : 0.058] | [-16.8 : 22.8] | [-7.7 : 9.3] | [-14.1 : 12.5] |
|  |  |  | Limit of Agreement Interval width | 0.106 | 0.074 | 39.6 | 17.0 | 26.6 |
|  |  |  | p | <0.001 | <0.001 | <0.01 | 0.09 | 0.34 |

R<sup>2</sup>, measurement of the goodness of fit of a model; RMSE, root mean square error of the linear regression fit; Slope, slope of the regression equation; Intercept, intercept of the regression equation; Interval of agreement 95%, the Interval of agreement at 95%; Limit of Agreement Interval width, width of the interval of agreement at 95%; p, probability of paired t-test for mean difference = 0. aBMD, areal Bone Mineral Density; vBMD, volumetric Bone Mineral Density; sBMD, surface Bone Mineral Density.

**Figure 1.**
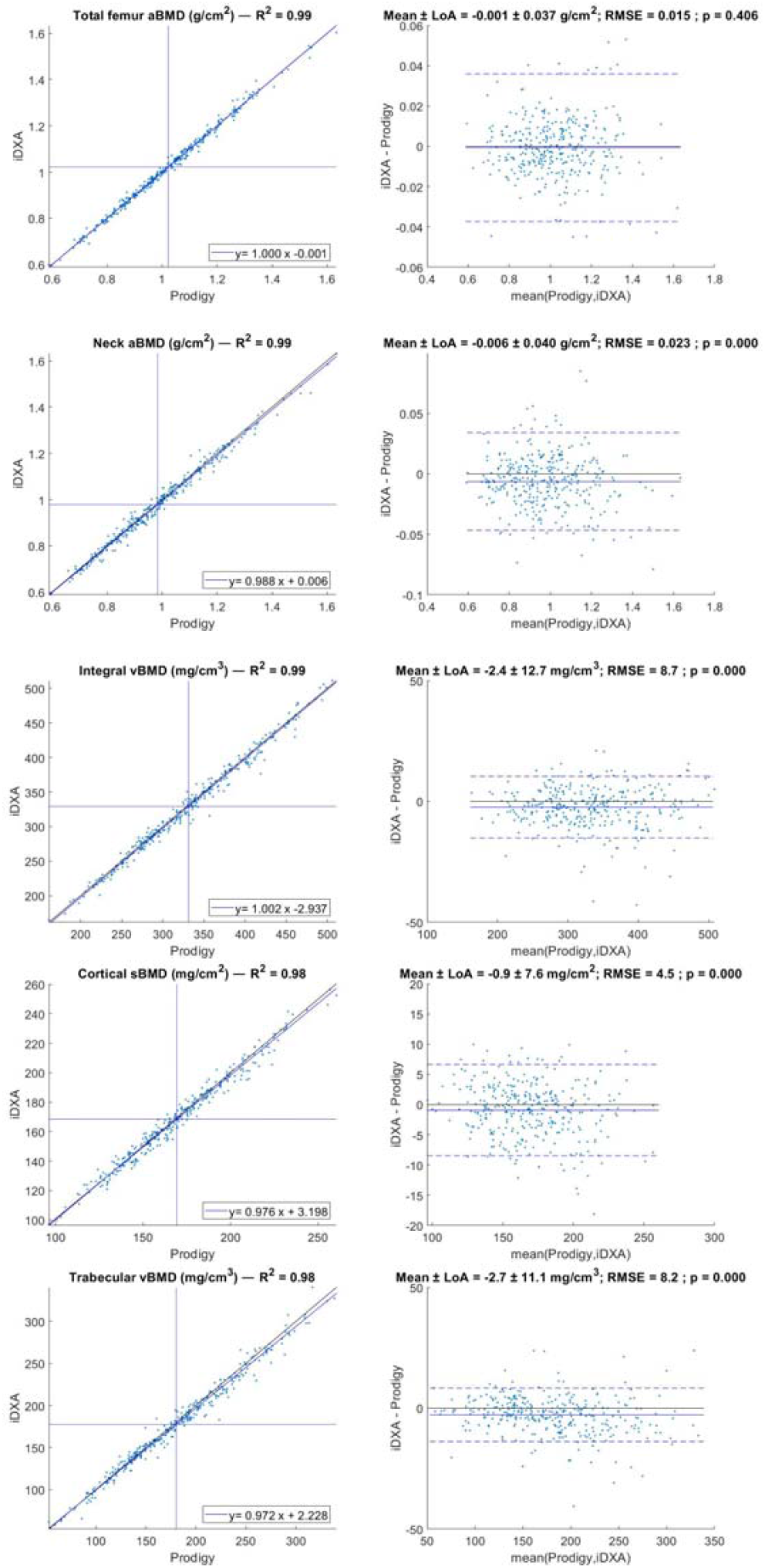
Deming regression (left) & Bland-Altman plot (right) between Prodigy and iDXA devices at University of Wisconsin for aBMD and 3D-Shaper parameters. In Bland-Altman plot, the blue line represents the bias while the dotted blue lines represent the LOA interval at 95%. aBMD, areal Bone Mineral Density; R^2^, measurement of the goodness of fit; Mean, bias between the two devices; LoA, Limit of Agreement; RMSE, Root Mean Square Error; p-value, probability of paired t-test for mean difference=0.

**Figure 2.**
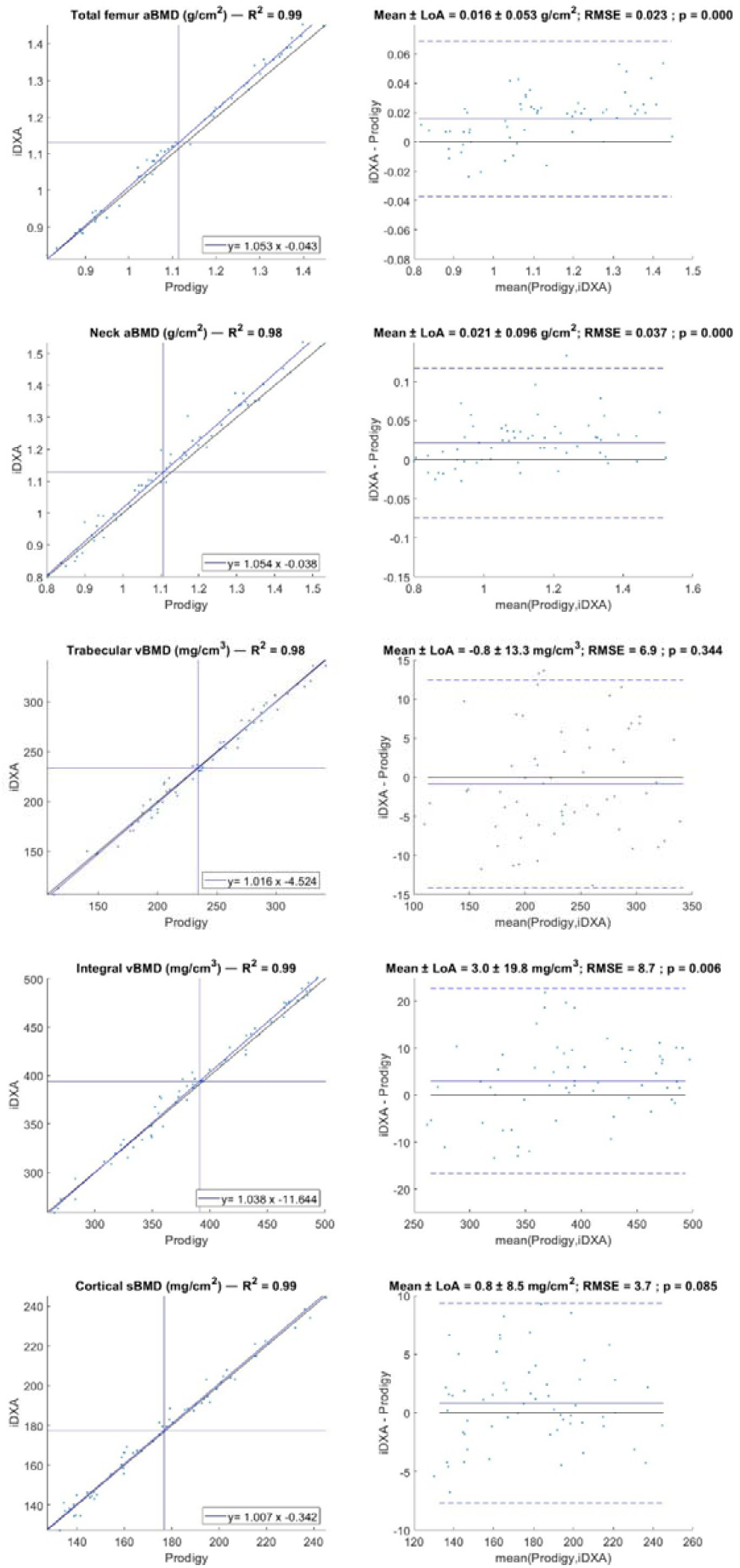
Deming regression (left) & Bland-Altman plot (right) between Prodigy and iDXA devices at California State University San Bernardino for aBMD and 3D-SHAPER parameters. In Bland-Altman plot, the blue line represents the bias while the dotted blue lines represent the LOA interval at 95%. aBMD, areal Bone Mineral Density; R^2^, measurement of the goodness of fit; Mean, bias between the two devices; LoA, Limit of Agreement; RMSE, Root Mean Square Error; p-value, probability of paired t-test for mean difference=0.

**Figure 3.**
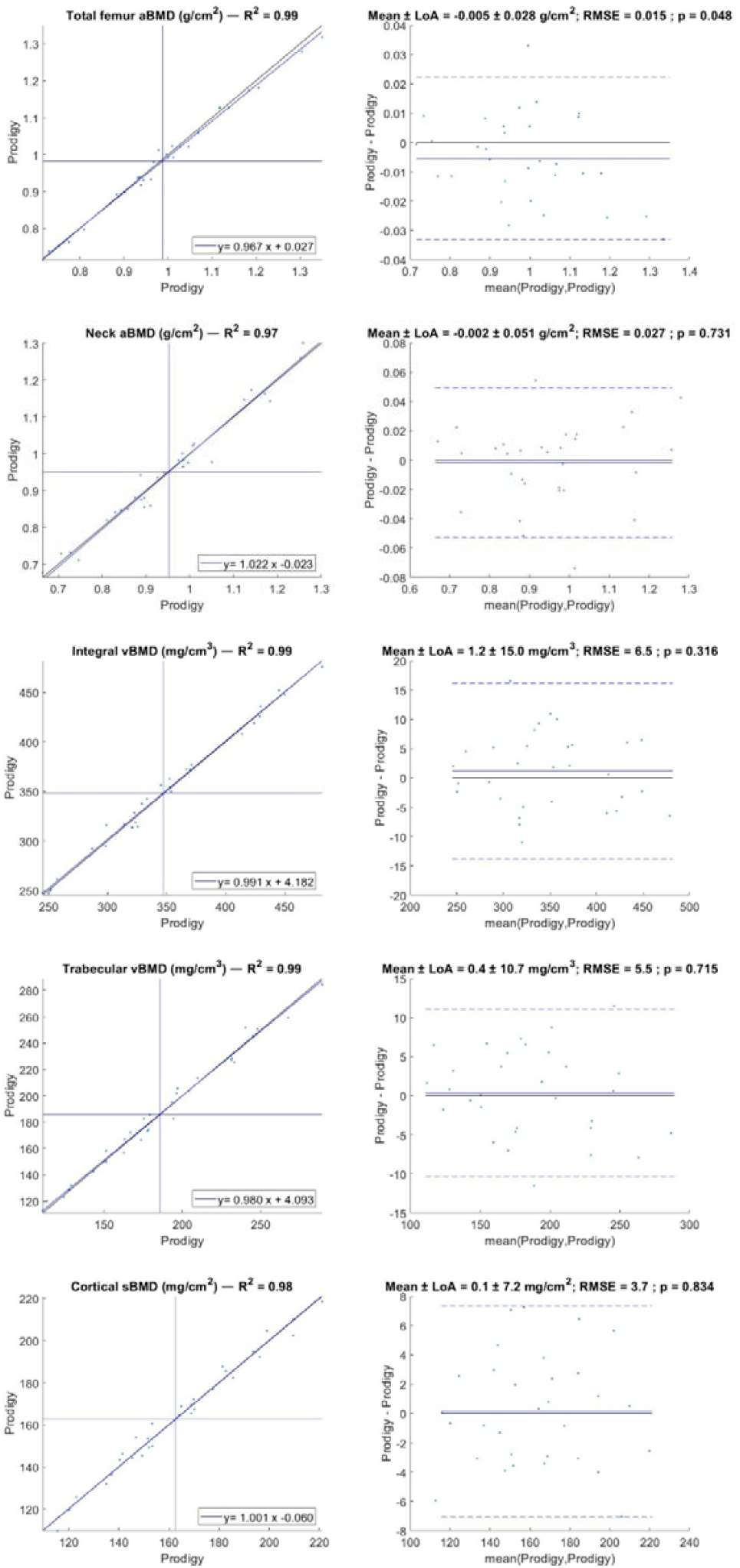
Deming regression (left) & Bland-Altman plot (right) between Prodigy and iDXA devices at Federal University of Rio de Janeiro for aBMD and 3D-SHAPER parameters. In Bland-Altman plot, the blue line represents the bias while the dotted blue lines represent the LOA interval at 95%. aBMD, areal Bone Mineral Density; R^2^, measurement of the goodness of fit; Mean, bias between the two devices; LoA, Limit of Agreement; RMSE, Root Mean Square Error; p-value, probability of paired t-test for mean difference=0.

#### 3D-DXA

Strong agreements between DXA scanner pairs at the respective sites (Table 2 and Figure 1,2 and 3) were obtained for integral vBMD, trabecular vBMD and cortical sBMD regardless of the center. Measurements were highly correlated as characterized by a R^2^ ranging from 0.98 to 0.99. In all centers, biases (in absolute value) were lower than 3.0 mg/cm^3^ for Integral vBMD, 2.7 mg/cm^3^ for trabecular vBMD and 0.9 mg/cm^2^ for cortical sBMD. Significant biases were obtained for integral vBMD, trabecular vBMD and cortical sBMD between Prodigy and iDXA for the UW (p<0.001) while significant bias was obtained only for integral vBMD (p<0.01) between Prodigy and iDXA at CSUSB. No significant biases (p ranging from 0.32 to 0.83) were demonstrated at FURJ between Prodigy devices. Similarly, no significant biases were observed for cortical sBMD (p=0.09) or trabecular vBMD (p=0.34) between Prodigy and iDXA at CSUSB (Table 2). Similar RMSE were obtained across centers, no significance by F-test (data not shown), for integral vBMD, cortical sBMD or trabecular vBMD. Limits of agreement interval widths were similar for trabecular vBMD (ranging from 21.4 to 26.6 mg/cm^3^) and cortical sBMD (ranging from 14.4 to 17 mg/cm^2^) regardless of the center (Table 2). Considering integral vBMD, limits of agreement interval widths measured at UW (25.4 mg/cm^3^) and FURJ (30 mg/cm^2^) while slightly wider, compared to limits of agreement interval width measured at CSUSB (39.6 mg/cm^3^).

#### Short-term precision

Short-term precision results are presented in Tables 3. Precision of total hip and femoral neck aBMD measurements was numerically lower at UFRJ and CSUSB, compared to UW. At UW, similar precision was obtained for aBMD at the total hip (RMS-SD: 0.006 g/cm^2^ for both Prodigy and iDXA) and at the femoral neck (RMS-SD: 0.010 and 0.009 g/cm^2^ for Prodigy and iDXA, respectively). At UFRJ, similar precision was obtained for aBMD at the total hip (RMS-SD: 0.011 and 0.009 g/cm^2^ for Prodigy 1 and Prodigy 2 respectively) while lower precision was observed at femoral neck for Prodigy 1 (RMS-SD: 0.023 g/cm^2^) when compared to Prodigy 2 (RMS-SD: 0.014 g/cm^2^). At CSUSB, similar precisions were obtained for aBMD at total hip and at femoral neck for iDXA (RMS-SD: 0.010 and 0.014 g/cm^2^ respectively). Regardless of the center or the device, similar precision was obtained for trabecular vBMD (RMS-SD ranging from 3.073 to 3.909 g/cm^3^), for cortical sBMD (RMS-SD ranging from 2.116 to 2.392 g/cm^2^), while higher variations were observed for integral vBMD (RMS-SD ranging from 2.989 g/cm^3^ to 4.139 g/cm^3^). Precision of integral vBMD measured using Prodigy devices was numerically slightly lower (RMS-SD ranging from 3.590 to 4.139 g/cm^3^), compared to iDXA (RMS-SD: 2.989 g/cm^3^).

**Table 3.**
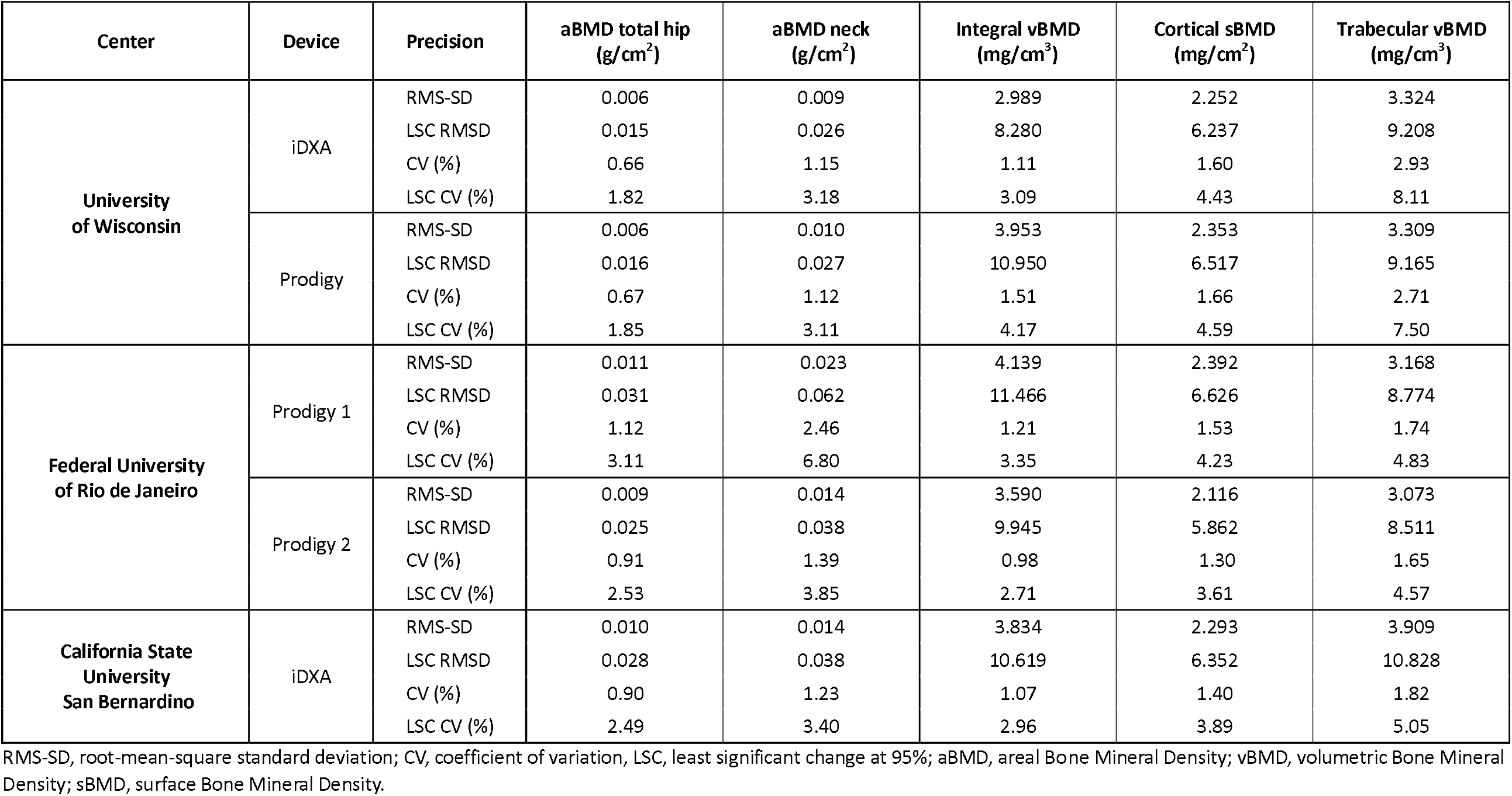
Precision of aBMD and 3D-DXA measurements by center.

## Discussion

This is the first study to evaluate the agreement between different GE Healthcare DXA scanners for 3D-DXA parameters. Strong agreements were observed between devices for both aBMD and 3D-DXA parameters. The aBMD agreements in this study are comparable to those previously reported in the literature [8, 9]. Results also showed similar RMSE values (all F-tests non-significant) across the centers, suggesting similar behavior of aBMD and 3D-DXA measurements between devices, regardless of center.

Bland-Altman analysis (Figures 1 to 3 and Table 2) was used to further evaluate the inter-scanner variations. Biases reported for total hip and neck aBMD, although statistically significant for some centers, were much lower than least significant changes assessed in this study for Prodigy and iDXA scanners, or those reported in the literature for such devices [8, 9], suggesting no clinical relevance. When evaluating 3D-DXA measurements, statistically significant biases were obtained at UW and at CSUSB for integral vBMD only. Biases reported for 3D-DXA measurements were lower than one third of the least significant change obtained in this study (Table 3), again making clinical impact negligible. Finally, the absolute slope differences from unity were lower than 6.0% for aBMD parameters (Table 2) and lower than 4.0% for 3D-DXA measurements (Table 2), showing strong agreement across DXA scanners. All together, these results support that aBMD and 3D-DXA parameters are measured similarly by Prodigy and iDXA densitometers.

In this study, aBMD short-term precision ranged from good (at FURJ) to excellent (at UW and CSUSB) on both Prodigy and iDXA densitometers, with CV% ≤ 1.12% at the total hip and CV% ≤ 2.46% at the femoral neck. These values are similar to those often reported in the literature [8, 9] and lower than the minimum acceptable precision for an individual technologist (CV% = 1.8% at the total hip and CV% = 2.5% at the femoral neck) [15]. The discrepancies observed across the centers participating in the current study could be explained by technical factors including variation of sample age, measurement stability of the device during and consistency of patient positioning or analysis performed by the technician.

Integral vBMD and total hip aBMD, which both measure the integral density at the total hip, showed similar precision. Furthermore, 3D-DXA parameters in this study demonstrated similar to, or better, precision than that previously reported for the iDXA device in earlier work [11], which documented RMS-SD of 3.75 mg/cm3, 2.28 mg/cm^2^ and 3.48 mg/cm^3^ for integral vBMD, cortical sBMD and trabecular vBMD, respectively, compared to 2.99 mg/cm^3^, 2.25 mg/cm^2^ and 3.32 mg/cm^3^ in the present study. Finally, similar precision was obtained for 3D-DXA parameters across all these DXA devices, including one Prodigy and one iDXA device at UW, two Prodigy devices at FURJ and one iDXA device at CSUSB (Table 3). Furthermore, it should be noted that the precision of 3D-DXA measurements was more consistent across the different scanners evaluated than precision error of aBMD measurements. For instance, the precision of total hip aBMD measurements at FURJ (RMS-SD = 0.009-0.011 g/cm^2^) and at CSUSB (RMS-SD = 0.010 g/cm^2^) were about 66% lower than at UW (RMS-SD = 0.006 g/cm^2^). The precision of 3D-DXA measurements remained quite stable across the different densitometers at FURJ and UW with RMS-SD ranging from 2.116 g/cm^2^ to 2.392 g/cm^2^ for cortical sBMD and from 3.073 g/cm^3^ to 3.909 g/cm^3^ for trabecular vBMD. Larger differences in precision results were observed between CSUSB and UW for integral vBMD (+22% at CSUSB) and for trabecular vBMD (+15% at CSUSB) on iDXA. However, these differences were lower than those obtained for aBMD at total hip (+66% at CSUSB) or at femoral neck (+40% at CSUSB). Interestingly, the lower precision of aBMD measurements at FURJ did not negatively affect the precision of 3D-DXA measurements at this center.

Such consistent precision across scanners may be explained by the 3D-Shaper modeling process, which estimates and corrects for the proximal femur’s position and orientation before automatically defining regions of interest and calculating 3D density parameters. Consequently, 3D-derived density measures should be less affected by patient positioning than conventional 2D DXA-based measurements. Moreover, the regions of interest for 3D-DXA parameters are automatically and consistently defined on a personalized 3D surface mesh generated from each patient’s DXA scan, minimizing technician-dependent variability. This reduced dependence on operator technique and scanning center could simplify precision assessments in densitometry facilities. In addition, because 3D-DXA parameters are less sensitive to patient positioning, they may also streamline positioning protocols, which are often time-consuming and challenging to implement in elderly populations.

The cohort used in this study consisted of 391 subjects covering a wide range of profiles. This combined cohort included both genders with varying anthropo-morphometric characteristics, age ranged from 20 to 90 years, height from 134 to 198 cm, weight from 47 to 136 kg and BMI from 17.4 to 48.8 kg/m^2^. Subjects from the UW and FURJ are representative of patient profiles commonly referred for DXA bone health assessment in routine clinical practice while the cohort from CSUSB is a homogenous group composed of young athletes. Similar inter-scanner variabilities were obtained in this study regardless of the center and study groups. Consequently, the results obtained in this study are unlikely to be related to homogeneous patient samples but more to technical difference existing between the devices.

Limitations of this study include performance in clinical research settings performed at only three centers using Prodigy and iDXA devices exclusively. Therefore, these results may not be generalized to other DXA devices pairs, manufacturers, patient populations or technologists. It should be noted that inter-device measurement performance can be influenced by the precision and accuracy of both the DXA device and 3D-Shaper reconstruction, as well as positioning and analysis by the technologist and patient population characteristics. In this study, the small overall errors observed suggest strong robustness and consistency of both aBMD and 3D-DXA measurements. Another limitation that precision assessments were conducted primarily in postmenopausal women, which may limit the applicability of these results to younger individuals and men.

## Conclusion

In conclusion, this is the first study evaluating inter-scanner variability for 3D-DXA measurements. Excellent agreement and comparable precision were observed between Prodigy and iDXA densitometers, as well as between Prodigy devices. Although statistically significant biases were observed across scanners, they were less than one-third of the LSCs, making clinical impacts negligible. The findings also demonstrate that no adjustment is required when using 3D-Shaper acquired on iDXA and Prodigy instruments.

## Data Availability

All data produced in the present work are contained in the manuscript.

## Author contributions

**Diane Krueger:** Conceptualization, Methodology, Formal analysis, Investigation, Writing - Review & Editing; **Neil Binkley:** Conceptualization, Methodology, Investigation, Resources, Writing - Review & Editing; **Miguel Madeira:** Investigation, Resources, Writing - Review & Editing; **Zhaojing Chen:** Investigation, Resources, Writing - Review & Editing; **Silvana Di Gregorio:** Investigation, Resources, Writing - Review & Editing; **Luis Miguel Del Río Barquero**: Investigation, Resources, Writing - Review & Editing; **Ludovic Humbert:** Conceptualization, Formal analysis, Writing - Review & Editing.

## Acknowledgment

We would like to thank Renaud Winzenrieth for his assistance in the First draft writing, editing and proofreading of this manuscript. We are also grateful to the volunteers and patients who participated in this research and made this work possible.

